# Liver discard rate due to conservative estimations of steatosis: an inference-based approach

**DOI:** 10.1101/2023.12.04.23299406

**Authors:** Hao Guo, Boris L. Gala-Lopez, Ian P.J. Alwayn, Kevin C. Hewitt

**Author notes:** Correspondence information: Kevin C. Hewitt, 6310 Coburg Road, PO Box 15000, Halifax, NS Canada B3H 4R2. Each author’s specific contributions to the work: H.G. conceived of the study and its design; acquisition, analysis, and interpretation of data, drafting and critically revising the manuscript for important intellectual content. K.C.H. provided oversight of the work in their supervisory capacity of the trainee H.G., K.C.H., B.L.G-L. and I.P.J.A. were involved in the critical revision of the manuscript. All authors approved the final version to be published.

## Abstract

**Background:** On-site conservative estimations of steatosis could result in the unnecessary discard of donor livers. This study applied the body mass index (BMI) as an independent statistical indicator to determine the extent of this problem. We explored two hypotheses: (I) that because of varying levels of expertise and protocols (reputational risk for pathologists), biopsies at transplant centers overestimate hepatic steatosis (HS), and (ii) that non-biopsy donor liver assessments are more conservative than biopsy-based evaluations.

**Methods:** The study processed cross-database and intra-database comparisons using data from the National Health and Nutrition Examination Survey (NHANES) and Organ Procurement and Transplantation Network (OPTN) spanning January 2017 to March 2020 in the United States. Post-matching BMI was applied as an independent indicator of statistical risk of HS.

**Results:** *Contrary to our first hypothesis*, biopsies at transplant centers did **not** overestimate HS - biopsy-classified donor livers were found in similar or lower risk categories. *Consistent with our second hypothesis*, absent biopsies, evaluations before and during organ procurement **were** observed to be more conservative, leading to the discard of 11.9% (373) of potential donor livers.

**Conclusions:** The study concludes that there was a significant (11.9%) disparity caused by on-site non-biopsy assessments of HS, leading to the unnecessary discard of potential donor livers. The findings emphasize the need to develop more accurate intraoperative techniques for assessing HS to optimize donor liver procurement.

## 1 Introduction

Hepatic steatosis (HS), especially moderate and severe macrovesicular steatosis (MaS) of donor livers, is associated with an increased risk of graft dysfunction after liver transplantation (LT).^1–4^ Although marginal donor livers have been accepted more and more often by transplant centers, HS remains a critical reason for discard.^5^ Livers with severe MaS (≥60%) are usually considered high risk for graft dysfunction and are often discarded outright.^6^ Livers with microvesicular steatosis (MiS) and mild (5%-30%) MaS are usually safe for LT, but transplanting livers with moderate (30-60%) MaS is still controversial.^6–9^ Furthermore, there seems to be disagreement between transplant physicians on the exact risk attributed to this steatosis. As a result, surgeons from different centers act differently when offered a steatotic organ.^10^ Therefore, accurate classification and quantification of steatotic donor livers are essential.

The current qualification of HS of donor livers relies on semi-subjective and subjective methods.^11^ Liver biopsy, though with limitations such as invasiveness, sampling errors, inter-observer variability, weak reproducibility, and occasional low on-site availability, is currently the “gold standard” for assessing HS of donor’s livers.^7,12,13^ However, a biopsy is only performed in a portion of donor surgeries. The most widely used method is the surgeon’s visual quality inspection during organ procurement surgeries. An online survey indicated that one-third of surgeon members of the American Society of Transplant Surgeons replied that visual inspection\ played a critical role in their decision to transplant more than 50% of donor livers.^14^ In contrast with visual estimation, only 30% of donors after cardiac death had data from biopsies between 2004 and 2010 in the United States.^3^

Surgeon’s visual quality inspection is based on the stiffness and macroscopic appearance of livers during organ procurement surgeries. Experienced surgeons can accurately predict <30% MaS; however, surgeons’ prediction of ≥□30% MaS was correct only for 52.2% of the cases, according to a prospective, double-blind study of 201 procured donor livers.^15^ Compared with the visual quality inspection, the degree of HS in biopsies is assessed more quantitatively; however, it shows a considerable variation among assessments of pathologists.^13^ It is also reported that the morphological semiquantitative evaluation of the degree of HS was constantly overestimated in a study on 75 liver biopsies.^16^

The low accuracy and high variability of the degree of HS assessed by the visual inspection and biopsies bring uncertainties to transplanting livers with MaS ranging 20%-60%. A natural hypothesis is that to ensure LT’s safety, transplant centers tend to estimate the degree of HS and make the decision of LT using a **conservative** approach, especially in cases without the availability of liver biopsies. Assuming this hypothesis is true, a considerable percentage of donor livers with “actual” MaS<30% are rejected before and/or during organ procurement. Since the “actual” degree of HS cannot be determined using the current gold standard, an alternative method is needed to quantify the discrepancies between the prevalence of HS and the frequency of HS of donor livers based on on-site assessments.

Since the information on the degree of HS is unavailable from donors who did not receive liver biopsies, the body mass index (BMI) can be used as an indirect intermediate parameter to statistically describe “potential” HS in each group of analyses. The risks of HS have been found significantly and independently correlated with increasing BMI in a nonlinear fashion.^17–19^ BMI was also found an independent predictor of the extent of fat infiltration.^20^ As a predictor for HS, the receiver operating characteristic (ROC) curve analysis of BMI were reported with an area under the receiver operator characteristic curve (AUROC) ranging from 0.77 to 0.84.^21,22^

In the present study, by adopting BMI as an independent indicator of the statistical HS risk, we applied an inference-based approach to quantify unrecovered steatotic donor livers impacted by on-site conservative estimations of biopsy and non-biopsy assessments. The analysis is supported by data from publicly available national clinical registries.

## 2 Materials and Methods

We analyzed the National Health and Nutrition Examination Survey (NHANES) 2017-March 2020 pre-pandemic data and the Organ Procurement and Transplantation Network (OPTN) data in the same timeframe in the United States. This research does not require Research Ethics Board review because it relies exclusively on information that is publicly available through a mechanism set out by regulation and that is protected by law, according to the 2018 Tri-Council Policy Statement for the Ethical Conduct for Research Involving Humans.

Designed to assess the health and nutritional status of adults and children in the United States, the NHANES examines a nationally representative sample every year. The National Center for Health Statistics combined the 2019-March 2020 data (uncompleted collection in the 2019-2020 cycle) with the 2017-2018 data using additional weighting procedures to provide nationally representative estimates.^23^

Although NHANES is population-wide, whereas OPTN captures donors, the two registries are comparable. 97% of the donor deaths collected by OPTN in 2009-2020 were caused by anoxia, stroke, head trauma, or central nervous system tumor, the incidence of which are stochastic and independent from BMI or liver diseases.^24^ Considering that the online database developed by OPTN contains data about every organ donation and transplant event in the United States since 1987,^25^ the OPTN data can also be regarded as nationally representative.

All statistical data analyses using the NHANES and OPTN data (Table **1**) were preformed using Stata Version 17.0-Basic Edition (StataCorp LLC, TX, USA). P-values of less than 0.05 were considered statistically significant.

**Table 1.**
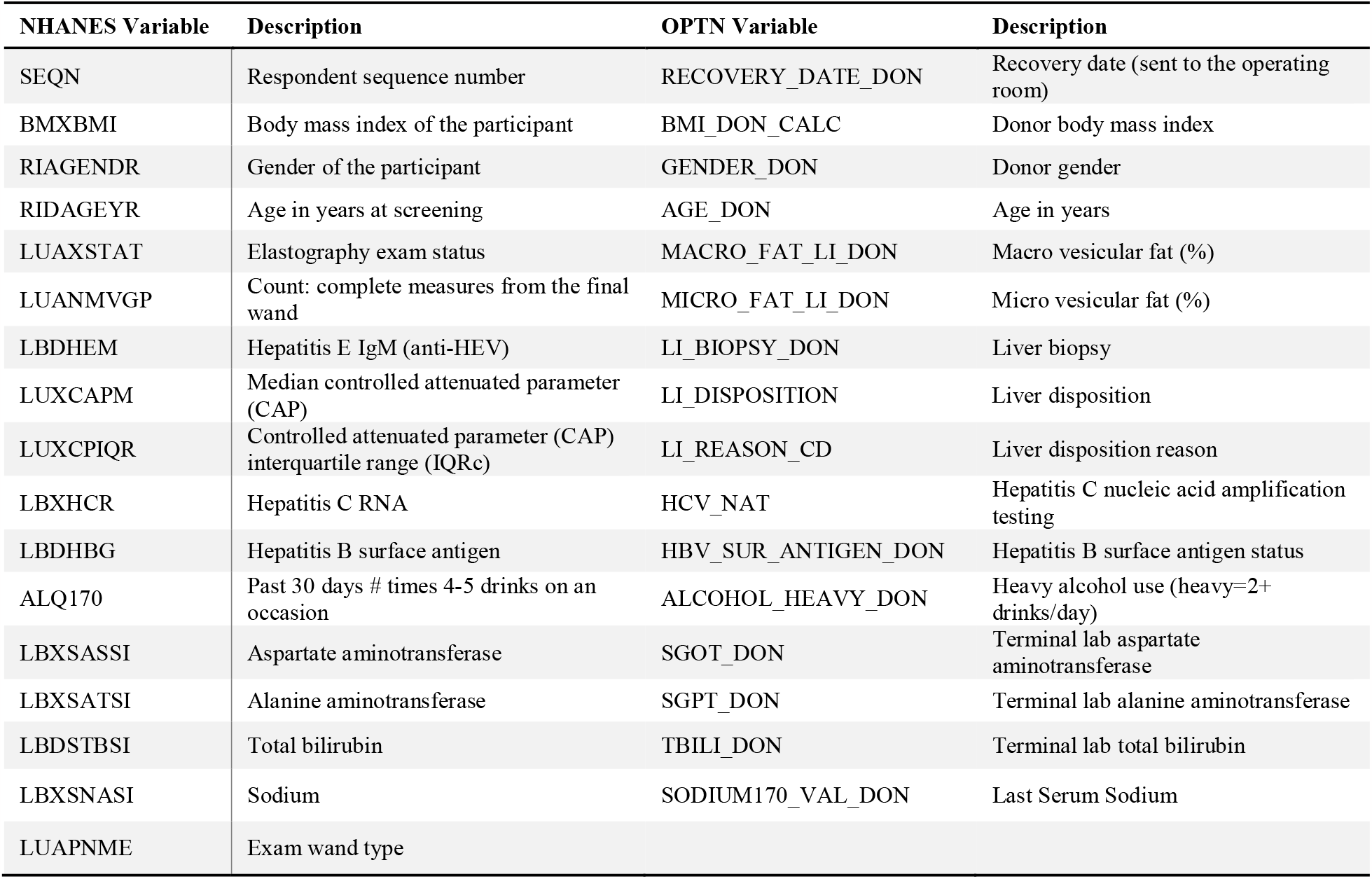
Variables of retrieved data from NHANES and OPTN registries.

### 2.1 The NHANES Data

Starting in 2017, liver ultrasound transient elastography measured by FibroScan^®^ was included in the NHANES. The controlled attenuation parameter (CAP) algorithm assesses ultrasonic attenuation and therefore examines the degree of HS. Following the Liver Ultrasound Transient Elastography Procedures Manuals, the FibroScan® model 502 V2 Touch equipped with a medium (M) or extra-large (XL) probe was used to obtain the elastography measurements and record the CAP measurements.^27,28^

Although many studies have shown good performance of CAP for excluding moderate and severe HS in potential living liver donors, the reported optimal cut-offs of CAP varied and could bring significant uncertainty in grading the degree of HS.^7,29–31^ To apply the most reasonable CAP cut-offs for this present cross-database study, we referred to a recent meta-analysis including 13 studies and 2,346 biopsy-controlled CAP datasets collected using the FibroScan^®^ M and XL probes^32^. In this present study, for data collected by the XL probes, the CAP cut-offs of non-HS (S_0_) versus mild HS (S_1_), S_1_ versus moderate HS (S_2_), and S_2_ versus severe HS (S_3_) were defined as 297 dB/m, 313 dB/m, and 333 dB/m, respectively.^32^ For data collected by the M probes, the CAP cut-offs of S_0_ versus S_1_, S_1_ versus S_2_, and S_2_ versus S_3_ were defined as 290.5 dB/m, 306.5 dB/m, and 326.5 dB/m, respectively, since the XL probes have been shown to negatively affect the CAP scores by 6.5 dB/m.^32^

### 2.2 The OPTN Data

In 2017-March 2020, the OPTN database recorded 36,014 deceased donors from whom at least one solid organ was recovered.

Deceased liver donations were classified into several categories for matching and analyses, as Table **2** describes. The MaS and HS cut-offs of S_0_ versus S_1_, S_1_ versus S_2_, and S_2_ versus S_3_ were defined as 5%, 30%, and 60%, respectively.

**Table 2.**
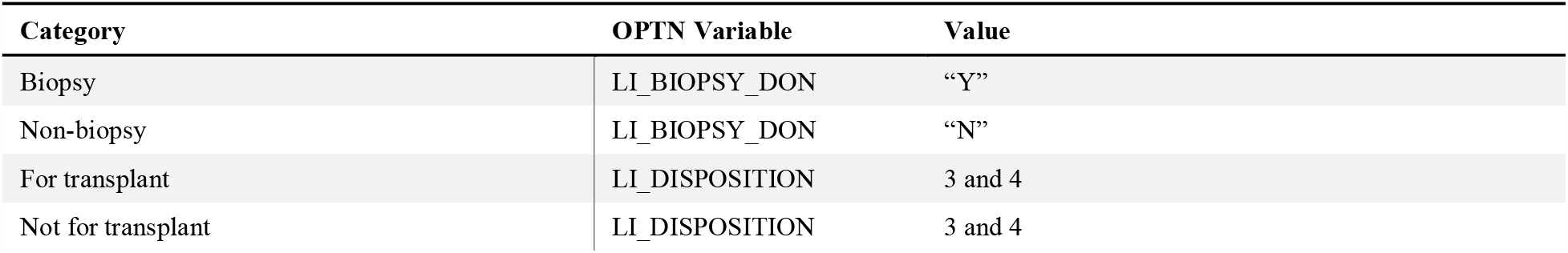
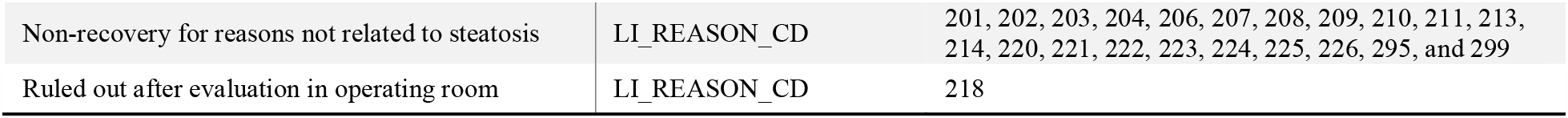
Classification of liver donations registered in OPTN.

| Category | OPTN Variable | Value |
| --- | --- | --- |
| Biopsy | LI_BIOPSY_DON | “Y” |
| Non-biopsy | LI_BIOPSY_DON | “N” |
| For transplant | LI_DISPOSITION | 3 and 4 |
| Not for transplant | LI_DISPOSITION | 3 and 4 |
| Non-recovery for reasons not related to steatosis | LI_REASON_CD | 201, 202, 203, 204, 206, 207, 208, 209, 210, 211, 213, 214, 220, 221, 222, 223, 224, 225, 226, 295, and 299 |
| Ruled out after evaluation in operating room | LI_REASON_CD | 218 |

### 2.3 Exclusion and Matching of Data

The included NHANES dataset consisted of 3,149 subjects (Figure **1**). Elastography measurements were included in the present study if (1) the examination contained at least ten complete measurements, (2) the participant was at least 18 years old, (3) the participant did not claim heavy alcohol use (defined as five or more alcoholic drinks for males or four or more alcoholic drinks for females on at least five days in the past month),^33^ (4) the participant did not carry viral hepatitis, and (5) the interquartile range of final CAP (IQRc) measures was non-zero and smaller than 40 dB/m^32,34,35^.

**Figure 1.**
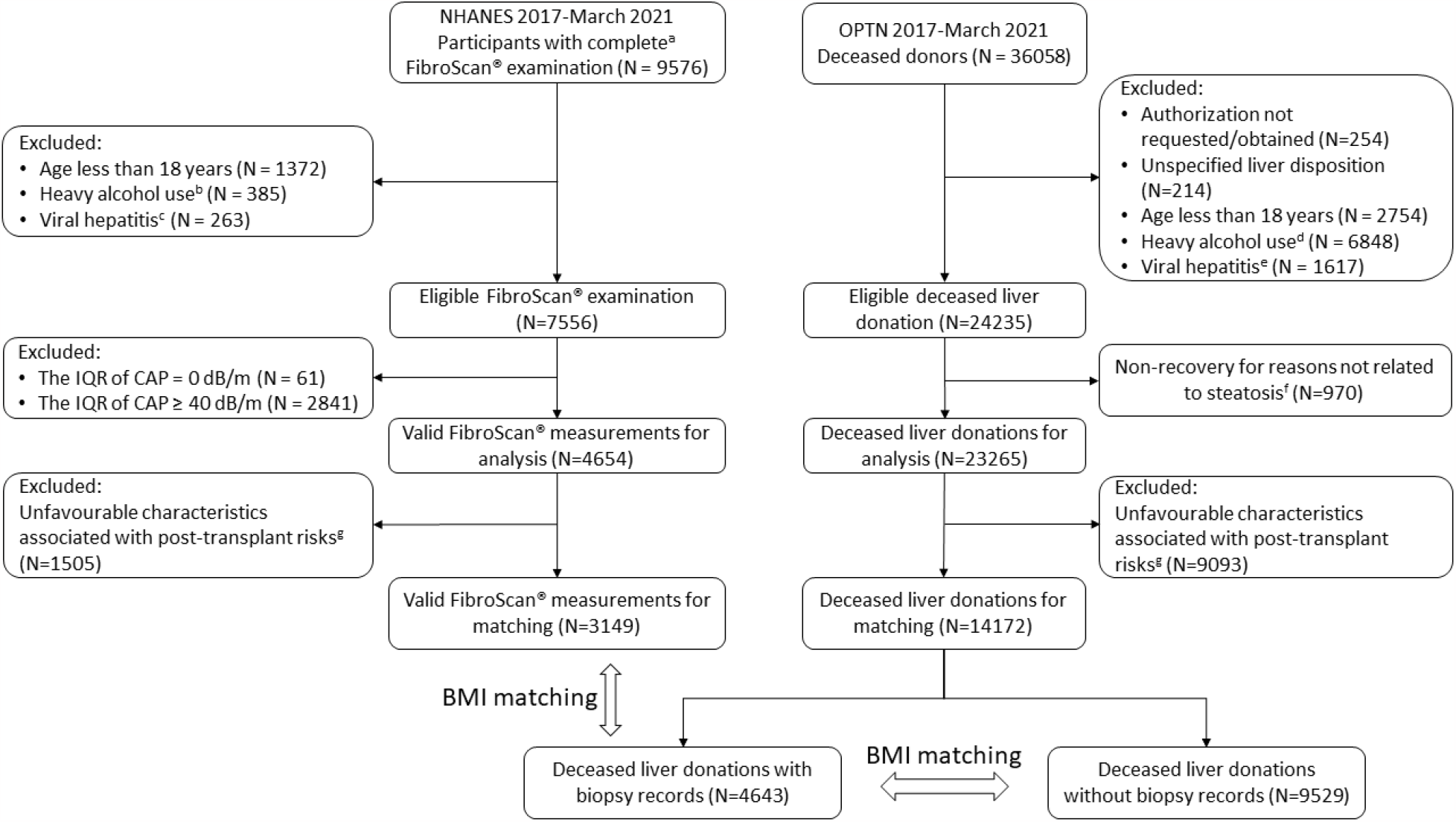
Flowchart of participants in the study of the prevalence of hepatic steatosis. (a) A complete FibroScan® examination contained at least ten complete measurements (the ratio interquartile range/median value of liver stiffness measurement (IQR/M) was not larger than 30%) of a patient fasting at least three hours. (b) Heavy alcohol use is defined as five or more alcoholic drinks for males or four or more alcoholic drinks for females on at least five days in the past month. (c) When the test result of any of hepatitis B surface antigen, hepatitis C RNA, hepatitis D antibody and hepatitis E IgM is positive, the participant was excluded. (d) The OPTN defines heavy alcohol use as consuming more than two drinks per day. (e) When the test result of any of hepatitis B surface antigen and hepatitis C RNA is positive, the donor was excluded. (f) The reasons not related to steatosis include cardiac arrest, positive hepatitis, positive HIV, anatomical abnormalities, vascular damage, no recipient located, donor medical history, donor social history, surgical damage in the operating room, hemodynamically unstable donor, trauma to the organ, time constraints, medical examiner restricted recovery, and other specify. (g) Participants and donors were excluded if at least one of the following criteria was met: age >65 years, serum sodium >165 mmol/L, ALT >105 U/L, AST >90 U/L, or serum bilirubin >3 mg/dl.

The included OPTN dataset consisted of 14,172 subjects (Figure **1**). Deceased liver donations were included in the present study if (1) an authorization of liver donation was obtained, (2) the donor was at least 18 years old, (3) the donor was not found to be a heavy alcohol user (defined as consuming more than two drinks per day),^36^ (4) the participant did not carry viral hepatitis, (5) the information of liver disposition was specified, and (6) the donor liver was either recovered, transplanted, or not recovered for reasons related to steatosis.

To eliminate potential interference of risk factors other than HS and BMI on this study, according to the definition of a liver donor marginal by European Association for the Study of the Liver (EASL) Clinical Practice Guidelines,^37^ elastography measurements and deceased liver donations were excluded from this study if any of the following criteria was met: age >65 years, serum sodium >165 mmol/L, alanine aminotransferase (ALT) >105 U/L, aspartate aminotransferase (AST) >90 U/L, or serum bilirubin >3 mg/dl.

In our study, we applied propensity score matching to match gender, age, BMI, ALT, AST, total bilirubin, and sodium of elastography participants (NHANES) and donors (OPTN) with biopsy records. As donor liver marginals had been excluded before matching, BMI was the only characteristic being matched for comparing donations (OPTN) with and without biopsy records. Propensity scores were calculated based on multivariable logistic regressions, where the dependent variable was the group identification (0 = group A, 1 = group B), and the independent variables were the characteristics to be matched.

Following the estimation of propensity scores, we used the “**psmatch2**” module in Stata to perform nearest-neighbor one-to-one matching.^38^ We assessed the quality of the matches by checking the balance of the covariates across the treated and untreated groups after matching using the “**pstest**” command.

## 3 Results

The pre- and post-matching population characteristics of included participants in the (NHANES) elastography study and donors registered in OPTN with biopsy data are shown in Table **3**, with the reporting of pre- and post-matching p-values. Records of 1,325 participants and 1,325 donors were matched. The post-matching participants and donors were well-correlated, as there was no statistically significant difference (p>0.05) in the characteristics, including age, BMI, ALT, AST, total bilirubin, and sodium. The post-matching data enabled comparison of prevalence of HS assessed by elastography and biopsy, among participants and donors with statistically same characteristics.

**Table 3.** Pre- and post-matching population characteristics of participants in the elastography study and donors registered in OPTN with biopsy records.

| Characteristics | Pre-matching |  |  | Post-matching |  |  |
| --- | --- | --- | --- | --- | --- | --- |
|  | NHANES | OPTN-Biopsy | P | NHANES | OPTN-Biopsy | P |
| Gender, N (%) |  |  |  |  |  |  |
| Female | 1685 (53.5) | 2327 (50.1) |  | 637 (46.9) | 637 (46.9) |  |
| Male | 1464 (46.5) | 2316 (49.9) |  | 688 (53.1) | 688 (53.1) |  |
| Age, yr, mean ± SD | | | | | | |
| Mean ± SD | 43.2 ± 14.1 | 47.2 ± 12.1 | <0.001 | 47.0 ± 13.7 | 46.8 ± 12.3 | 0.790 |
| Median (IQR) | 44 (31-56) | 49 (39-57) |  | 50 (37-59) | 49 (38-57) |  |
| BMI, kg/m<sup>2</sup> |  |  |  |  |  |  |
| Mean ± SD | 29.9 ± 7.2 | 32.0 ± 8.2 | <0.001 | 30.4 ± 7.3 | 30.4 ± 7.8 | 0.911 |
| Median (IQR) | 28.8 (25.1-33.5) | 30.9 (26.0-36.5) |  | 29.1 (25.3-34.1) | 29.2 (24.5-34.8) |  |
| ALT, U/L, median (IQR) |  |  |  |  |  |  |
| Female | 15 (12-22) | 28 (18-46) | <0.001 | 19 (13-29) | 19 (13-29) | 0.651 |
| Male | 23 (17-32) | 29 (19-46) | <0.001 | 25 (18-35.5) | 23 (16-37) | 0.831 |
| AST, U/L, median (IQR) |  |  |  |  |  |  |
| Female | 17 (15-21) | 29 (20-45) | <0.001 | 20 (16-26) | 20 (15-27) | 0.766 |
| Male | 21 (17-26) | 31.5 (22-50) | <0.001 | 22 (19-29) | 22.5 (16-32) | 0.519 |
| Total Bilirubin, mg/dL, median (IQR) |  |  |  |  |  |  |
| Female | 0.3 (0.2-0.5) | 0.6 (0.4-0.8) | <0.001 | 0.4 (0.3-0.6) | 0.4 (0.3-0.6) | 0.845 |
| Male | 0.4 (0.3-0.6) | 0.7 (0.4-1.0) | <0.001 | 0.5 (0.4-0.7) | 0.5 (0.4-0.8) | 0.388 |
| Sodium, mg/dL, median (IQR) |  |  |  |  |  |  |
| Female | 140 (139-142) | 149 (144-154) | <0.001 | 142 (140-143) | 142 (138-146) | 0.063 |
| Male | 141 (139-142) | 149 (143.5-154) | <0.001 | 142 (140-143) | 142 (139-145) | 0.052 |

S<sub>0</sub>, non-steatosis; S<sub>1</sub>, mild steatosis; S<sub>2</sub>, moderate steatosis; S<sub>3</sub>, severe steatosis; SD, standard deviation; BMI, body mass index; ALT, alanine aminotransferase; AST, aspartate aminotransferase; IQR, interquartile range. The median CAP scores classify S<sub>0</sub>-S<sub>3</sub>. Null hypothesis: there is a statistically significant difference in the distribution of a covariate between the treated and control groups after matching.

In Figure **2**, a stacked column chart was used to compare the prevalence of HS in the post-matching groups. Considering moderate and severe steatosis as risky for transplantation, elastography classified the most (28%) participants (NHANES) into the “risky steatosis” category. Biopsy classified similar percentage (25%) of donors (OPTN) into the “risky steatosis” category based on the degree of overall HS, showing no overestimation which was due to the hypothesized conservative assessment of donor livers. Applying the degree of MaS (which is more clinically relevant to graft dysfunction) as the classifier, significantly less (13%) donors were classified into the “risky steatosis” category, indicating that biopsy can lessen unnecessary discarding of steatotic donor livers.

**Figure 2.**
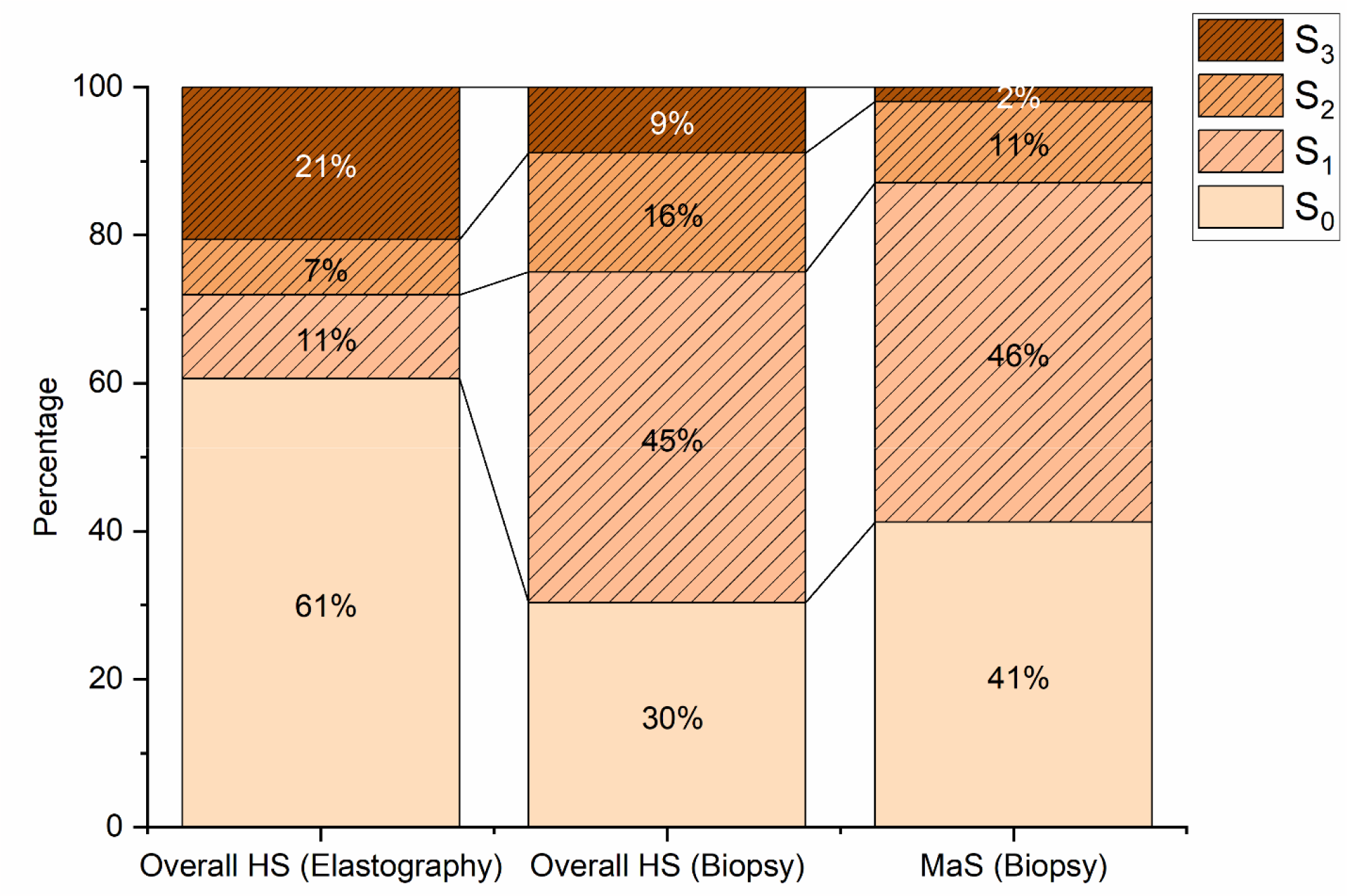
stacked column chart the prevalence of steatosis in the post-matching groups. S_0_, non-steatosis; S_1_, mild steatosis; S_2_, moderate steatosis; S_3_, severe steatosis. The median CAP scores and biopsy records classify S_0_-S_3_. Elastography classified more HS into S_0_ and S_3_, while biopsy classified less HS into the “risky” group S_2_ & S_3_

The elastography-based CAP score shows significant disagreement with biopsy in classifying HS and MaS in S_0_ and S_1_ stages. On condition that the post-matching population characteristics had no significant statistical difference, CAP scored classified the most elastography participants as non-steatotic (61%), whereas biopsies scored classified the most liver donors into mild overall HS (45%) or mild MaS (46%). The disagreement may support that biopsy is more conservative in classifying S_0_ and S_1_; however, as both S_0_ and S_1_ are considered “non-risky” in LT, this finding did not support that donor HS was assessed in a statistically conservative approach by biopsy.

The pre- and post-matching population characteristics of the included donors registered in OPTN with and without biopsy records are shown in Table **4**, with the reporting of pre- and post-matching p-values of the “pstest”. Since most donors (9,529, 67%) did not have biopsy records, only 4,264 donors without biopsy records were matched. The post-matching donors with and without biopsy records had no statistically significant difference (p>0.05) in the distribution of covariate between BMIs, indicating excellent BMI matching in this intra-group comparison. Although this less rigorous matching solely targeted BMI, the values of other risk factors, no matter their statistical differences were significant or not, were all within the safe ranges because the donations including unfavored characteristics had already been excluded before matching.

**Table 4.**
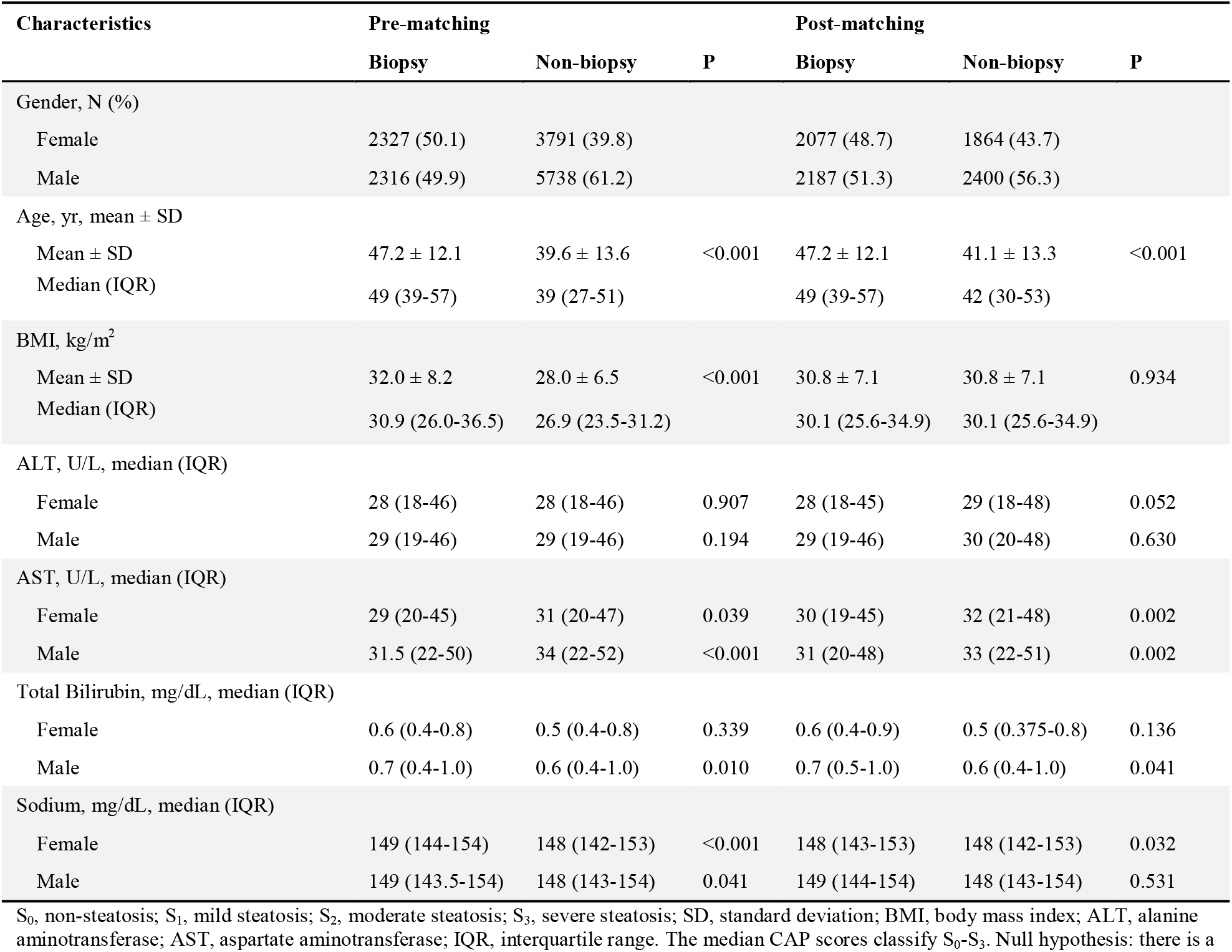
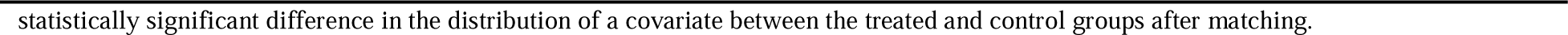
Pre- and post-matching population characteristics of donors registered in OPTN.

| Characteristics | Pre-matching |  | P | Post-matching |  | P |
| --- | --- | --- | --- | --- | --- | --- |
|  | Biopsy | Non-biopsy |  | Biopsy | Non-biopsy |  |
| Gender, N (%) |  |  |  |  |  |  |
| Female | 2327 (50.1) | 3791 (39.8) |  | 2077 (48.7) | 1864 (43.7) |  |
| Male | 2316 (49.9) | 5738 (61.2) |  | 2187 (51.3) | 2400 (56.3) |  |
| Age, yr, mean ± SD | | | | | | |
| Mean ± SD | 47.2 ± 12.1 | 39.6 ± 13.6 | <0.001 | 47.2 ± 12.1 | 41.1 ± 13.3 | <0.001 |
| Median (IQR) | 49 (39-57) | 39 (27-51) |  | 49 (39-57) | 42 (30-53) |  |
| BMI, kg/m<sup>2</sup> |  |  |  |  |  |  |
| Mean ± SD | 32.0 ± 8.2 | 28.0 ± 6.5 | <0.001 | 30.8 ± 7.1 | 30.8 ± 7.1 | 0.934 |
| Median (IQR) | 30.9 (26.0-36.5) | 26.9 (23.5-31.2) |  | 30.1 (25.6-34.9) | 30.1 (25.6-34.9) |  |
| ALT, U/L, median (IQR) |  |  |  |  |  |  |
| Female | 28 (18-46) | 28 (18-46) | 0.907 | 28 (18-45) | 29 (18-48) | 0.052 |
| Male | 29 (19-46) | 29 (19-46) | 0.194 | 29 (19-46) | 30 (20-48) | 0.630 |
| AST, U/L, median (IQR) |  |  |  |  |  |  |
| Female | 29 (20-45) | 31 (20-47) | 0.039 | 30 (19-45) | 32 (21-48) | 0.002 |
| Male | 31.5 (22-50) | 34 (22-52) | <0.001 | 31 (20-48) | 33 (22-51) | 0.002 |
| Total Bilirubin, mg/dL, median (IQR) |  |  |  |  |  |  |
| Female | 0.6 (0.4-0.8) | 0.5 (0.4-0.8) | 0.339 | 0.6 (0.4-0.9) | 0.5 (0.375-0.8) | 0.136 |
| Male | 0.7 (0.4-1.0) | 0.6 (0.4-1.0) | 0.010 | 0.7 (0.5-1.0) | 0.6 (0.4-1.0) | 0.041 |
| Sodium, mg/dL, median (IQR) |  |  |  |  |  |  |
| Female | 149 (144-154) | 148 (142-153) | <0.001 | 148 (143-153) | 148 (142-153) | 0.032 |
| Male | 149 (143.5-154) | 148 (143-154) | 0.041 | 149 (144-154) | 148 (143-154) | 0.531 |

S<sub>0</sub>, non-steatosis; S<sub>1</sub>, mild steatosis; S<sub>2</sub>, moderate steatosis; S<sub>3</sub>, severe steatosis; SD, standard deviation; BMI, body mass index; ALT, alanine aminotransferase; AST, aspartate aminotransferase; IQR, interquartile range. The median CAP scores classify S<sub>0</sub>-S<sub>3</sub>. Null hypothesis: there is a

Figure **3(a)** compares the distributions of post-matching BMIs between the groups of all donors with and without biopsy records. The kernel density curves almost fully overlap each other, which confirms that inherent discrepancy of BMI distributions does not impact the comparisons between donors with and without biopsy records in the following sub-groups.

**Figure 3.**
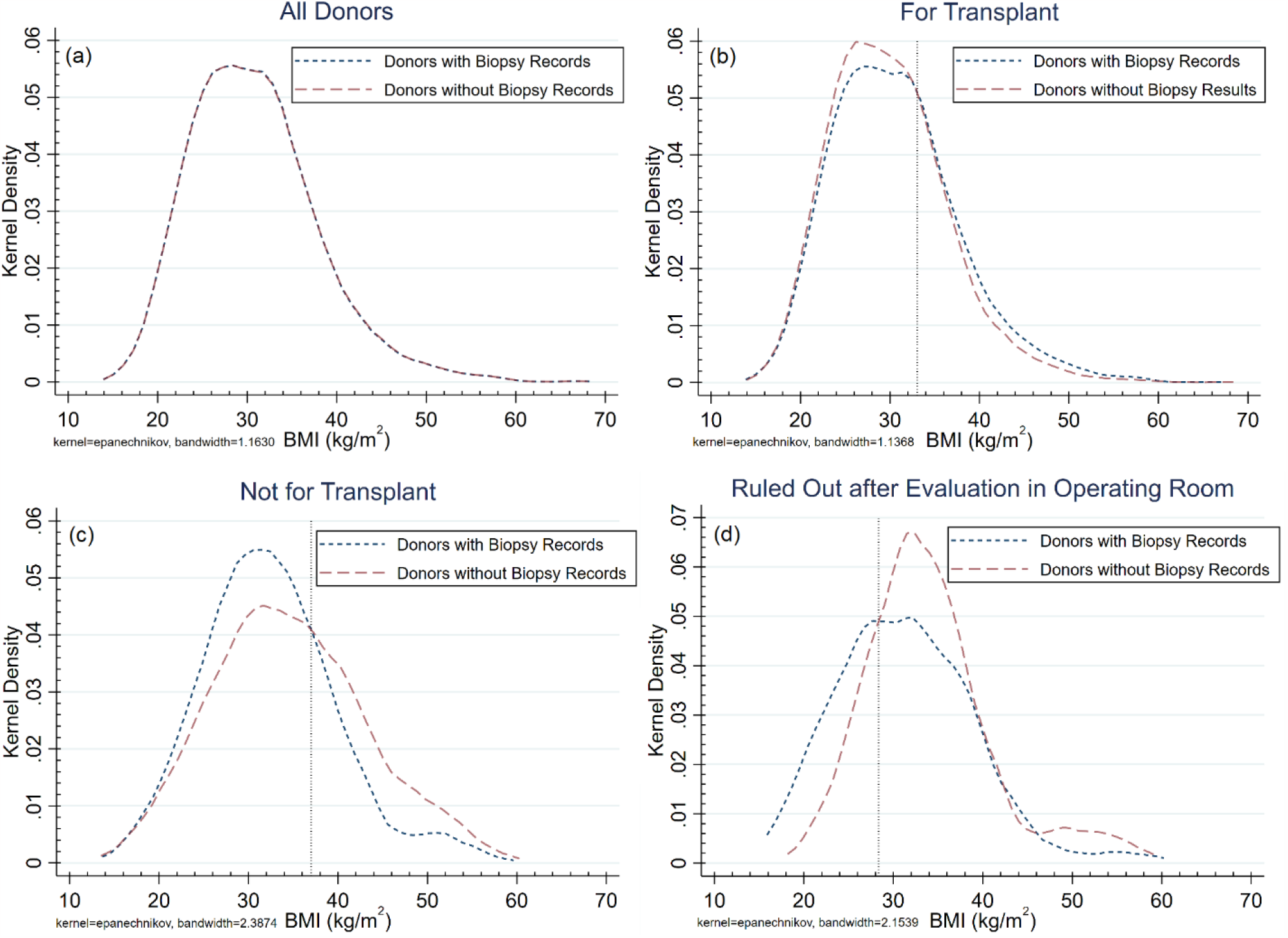
Univariate kernel density estimations of post-matching BMIs among subjects of different groups. (a) Comparison of BMIs between the group donors with and without biopsy records. (b) Comparison of BMIs between the group of donors without biopsy records and those with biopsy records in the group “For Transplantation.” The vertical dashed line at 32.5 kg/m indicated the corresponding BMI value at the cross point of the kernel density curves. (c) Comparison of BMIs between the group of donors without biopsy records and those with biopsy records in the group “Not for Transplant.” The vertical dashed line at 37.0 kg/m indicated the corresponding BMI value at the cross point of the kernel density curves. (d) Comparison of BMIs between the group of donors without biopsy records and ruled out after evaluation in the operating room and those with biopsy records ruled out after evaluation in the operating room. The vertical dashed line at 28.4 kg/m indicated the corresponding BMI value at the lower cross point of the kernel density curves.

Figures **3(b)** shows a comparison of the distributions of post-matching BMIs between the group of donors with biopsy records (N=3,929) and those without biopsy (N=3,421) records in the group “For transplant.” Among the donors livers “for transplant”, those with biopsy records were more frequent than those without biopsy records in the high-BMI region (where BMI is higher than the cross point at 32.5 kg/m^2^). 4.4% donor livers with biopsy records similarly high statistical risks of HS were precured for transplant (the area difference under the curves on the right side of the dashed line).

Meanwhile, among the discarded donor livers, those in the high-BMI region (where BMI is higher than the cross point at 37.0 kg/m^2^) were more likely without biopsy records. Figures **3(c)** shows comparisons of the distributions of post-matching BMIs between the group of donors with biopsy records (N=335) and those without biopsy (N=843) records in groups “Not for transplant.” 11.9% of donor livers “Not for transplant” with similarly high statistical risks of HS did not have biopsy data recorded. Converting the percentage numbers to the pre-matching OPTN records, 11.9% of 1,487 livers “not for transplant” without biopsy records (out of 9,529 liver donations without biopsy records for the data matching) led to 177 unrecovered donor livers. Enlarging the applied scope to the 24,365 eligible organ donors registered in the United States in 2017-March 2020, 11.9% of 3,407 livers “not for transplant” without biopsy records led to 373 unrecovered donor livers.

Figures **3(d)** compares the distributions of post-matching BMIs between the group of donors with biopsy records (N=142) and ruled out after evaluation in the operating room and the group of donors without biopsy records (N=140) and ruled out after evaluation in the operating room. Applying 28.4 kg/m^2^ (the corresponding BMI value at the lower cross point of the kernel density curves) as cut-off, comparing those with BMI lower than 28.4 kg/m^2^, donors ruled out after evaluation in the operating room and with BMI larger than 28.4 kg/m^2^ were 15.9% more frequent to be without biopsy results.

## 4 Discussions and Conclusion

This present study adopted BMI as an independent indicator of the statistical risk of HS and tested two hypotheses: (1) transplant centers tend to overestimate the degree of HS and make the decision of LT using a conservative biopsy, and (2) non-biopsy donor liver assessments are more conservative than biopsy-based assessments.

Tests on the first hypothesis were performed by comparing the population prevalence of HS in the United States in 2017-March 2020 and the prevalence of HS and MaS in American liver donors in the same timeframe. Potential interference of risk factors other than HS and BMI were excluded according to the definition of a liver donor marginal by EASL Clinical Practice Guidelines.^37^ To enable cross-database comparisons, rigorous data matching eliminated significant statistical differences between the NHANES population data of participants and the OPTN data of donors in gender, age, BMI, ALT, AST, total bilirubin, and sodium. Although biopsy classified less HS to the degree S_0_ and more to the degree S_1_, this finding did not support that transplant centers assessed donor HS in a statistically conservative approach by biopsy, as both S_0_ and S_1_ are considered “non-risky” in LT. On the contrary, 13% less donors were classified into the “risky steatosis” category according to the percentage of MaS, compared with classifications based on CAP scores.

The second hypothesis was evaluated utilizing a less stringent BMI matching strategy. Given that both the biopsy and non-biopsy data originated from the same OPTN registry, and that liver donors on the marginal fringe had been excluded prior to matching, the matching process was solely dependent on the donors’ BMI. As BMI is a positively correlated independent risk factor of HS,^17–19^ it was utilized as a surrogate to estimate the statistical risks of HS impacting the liver discard rates, specifically in the context of non-biopsy assessments of steatosis.

The analysis of post-matching OPTN data corroborated that on-site non-biopsy evaluations of donor livers tended to be more conservative than their biopsy-inclusive counterparts. Notably, 4.4% of donor livers with biopsy records exhibited comparably high statistical risks of HS but were still selected for transplantation. Conversely, among the discarded donor livers, biopsy data was absent for 11.9% of those presenting similar high statistical risks of HS. Translating these percentages into real numbers based on OPTN records, the conservative on-site non-biopsy assessment of HS inadvertently led to the loss of 177 to 373 donor livers in the United States from 2017 to March 2020. This suggests that an additional 177 to 373 livers could potentially have been recovered without surpassing the accepted risk threshold for HS.

A prominent disparity caused by on-site non-biopsy assessments HS could be observed in the operating room. Analysis of the post-matching BMI distributions between donors with and without biopsy records revealed that donors without biopsy records were discarded 15.9% more frequently compared to donors with biopsy records and comparable high BMIs.

While HS typically presents as a mixed form of MaS and MiS, the negative impact of MiS on post-LT dysfunction is significantly less pronounced than MaS.^39,40^ Therefore, determining HS in biopsied donor livers based on the degree of MaS has greater clinical relevance. However, the HS defined by CAP scores is not directly comparable to biopsy-defined MaS, despite the referenced meta-analysis being based on biopsy-controlled CAP datasets.^32^ Nevertheless, both overall HS (25%) and MaS (13%) assessments by biopsy classified fewer donor livers as “high-risk HS” compared to population prevalence (28%), effectively refuting the hypothesis that transplant centers tend to conduct overly conservative biopsies on donor livers.

A limitation of this study is that elastography measurements from NHANES were not biopsy-validated, although the CAP cut-offs were optimized using biopsy-controlled CAP datasets.

Although CAP cut-offs varied according to cause,^32^ we believe our utilized cut-offs were appropriate, given the exclusion of viral hepatitis and marginal donor livers. This assumption is indirectly supported by a 2020 histology-based study reporting a 38% prevalence of non-alcoholic fatty liver disease in a middle-aged US cohort,^41^ closely matching our 39% prevalence of CAP-based HS.

Another constraint is that to fulfill data matching criteria, most of the data (58%) remained untreated, which negatively affected the data volume. This study also purposefully omitted considerations of compound factors influencing donor liver quality. Although we excluded non-recoveries due to non-steatosis related reasons, livers classified as “not for Transplant” could be attributed to numerous factors, including poor quality of blood vessels, malignancies, fibrosis, technical procurement issues, and HS. However, these factors might not be thoroughly considered when extrapolating statistical HS risks to BMIs.

In this study, we translated comparisons between HS degrees in donor livers with and without biopsy records into comparisons of HS risks, using BMI as an indirect intermediate parameter to statistically depict HS risks. One advantage of this inferential approach is that it allowed us to quantify the impact of conservative on-site steatosis estimations on unrecovered donor livers on a statistical level. Nevertheless, the current research did not disclose how on-site assessments, including both biopsies and non-biopsy evaluations, impact liver recovery for individual donors. Individual case discrepancies might exceed statistical discrepancies due to uncertainties inherent in current subjective and semi-subjective HS assessment methods. Most donors (9,529, 67%) did not have biopsy records. Although the OPTN data did not enable us to track if a biopsy was operated before or during the organ procurement surgeries, the majority of the recorded biopsies were believed intraoperative biopsies, since most organ procurement organizations reported that 5% to 10% of their donors received a prerecovery liver biopsy, according to a study in 2017.^42^ The development of accurate and objective intraoperative techniques for assessing HS and MaS in the future will be vital to mitigate the impacts of conservative on-site estimations.

## Data Availability

All data produced are available online at:
https://wwwn.cdc.gov/nchs/nhanes/continuousnhanes/default.aspx?Cycle=2017-2020
https://optn.transplant.hrsa.gov/data/view-data-reports/request-data/

https://optn.transplant.hrsa.gov/data/view-data-reports/request-data/

## Abbreviations used in this paper

ALT: alanine aminotransferase
AST: aspartate aminotransferase
BMI: body mass index
CAP: controlled attenuation parameter
HS: hepatic steatosis
IQR: interquartile range
IQRc: interquartile range of final controlled attenuation parameter
LT: liver transplantation
M: medium
MaS: macrovesicular steatosis
MiS: microvesicular steatosis
NHANES: National Health and Nutrition Examination Survey
OPTN: Organ Procurement and Transplantation Network
SD: standard deviation
XL: extra-large

